# Fatigue in perinatal stroke is associated with resting-state functional connectivity

**DOI:** 10.1101/2020.04.14.20065326

**Authors:** J.G Wrightson, H.L Carlson, E.G Condliffe, A Kirton

## Abstract

Fatigue is prevalent in youth with perinatal stroke, but the causes are unclear. Predictive coding models of adult post-stroke fatigue suggest that fatigue may arise from dysfunction in predictive processing networks. To date, the association between fatigue and neural network connectivity in youth with perinatal stroke has not been examined. The present study examined the association between fatigue and the functional connectivity of predictive processing neural networks, measured using resting-state functional magnetic resonance imaging, in individuals with perinatal stroke. Participants who reported experiencing fatigue had weaker functional connectivity between the non-lesioned middle frontal and supramarginal gyri and between the non-lesioned intracalcarine cortex and the lesioned paracingulate cortex. In contrast, participants reporting fatigue had stronger functional connectivity between the lesioned inferior temporal gyrus and non-lesioned insula. These results suggest that fatigue in youth with hemiparetic cerebral palsy caused by perinatal stroke is associated with the functional connectivity of hubs previously associated with predictive processing and fatigue. These results suggest potential cortical and behavioral targets for the treatment of fatigue in individuals with perinatal stroke.

## Introduction

Perinatal stroke is the leading cause of hemiparetic cerebral palsy and results in lifelong physical disability ^1,2^. Fatigue is common in individuals with hemiparetic cerebral palsy, including those with perinatal stroke ^3–6^. Fatigue significantly reduces health-related quality of life in individuals with cerebral palsy ^7–9^, and reducing fatigue is caregivers’ highest priority ^10^. However, the mechanisms which give rise to fatigue are not well understood, and consequently, there are few evidence-based treatments.

Predictive coding models of fatigue suggest that insult to the central nervous system causes an incongruity between the efferent sensory predictions encoded by the brain’s generative model of the body and the afferent sensory prediction errors that arise from an action ^11,12^. It has been proposed that dysfunction in the predictive processing networks gives rise to fatigue ^11^. Predictive processing theoretical frameworks suggest that the brain contains a generative model of the body-state ^13^. Within these frameworks, fatigue arises from an ongoing and unresolvable discrepancy between the brain’s prediction of the body-state and the afferent sensory information signalling the actual-body state (the prediction) ^11,12,14^. These prediction errors signal a failure of the brain to predict the body state, i.e. an inability to maintain homeostasis. This ongoing discrepancy between predicted and actual body state is consciously perceived as the feeling of fatigue, which acts as a signal for allostatic behavioral adjustment: rest. Two predictive coding accounts of fatigue have been proposed; the first suggests that fatigue arises from a failure to attenuate from proprioceptive or exteroceptive sensory prediction errors, specifically during motor actions ^15^. An opposing account posits that fatigue arises through the metacognitive awareness of an ongoing discrepancy between predicted and actual interoceptive information, processed in an interoceptive predictive processing network ^12^. These two accounts are often presented as disparate, assigning fatigue to either interoceptive or exteroceptive and proprioceptive predictive processing (see Kuppuswamy 2021; Manjaly et al. 2019 for reviews). However, these systems likely interact to predict proprioceptive and interoceptive signals in the pursuit of goal-states ^16^ and thus are both likely to contribute to the perception of fatigue ^17^.

Unlike in adult stroke, where pre-stroke conditions and post-stroke responses are frequently assumed to contribute to the subsequent development of fatigue ^18,19^, fatigue in perinatal stroke is likely to be a consequence of the abnormal development of the brain ^6^. The interoceptive predictive processing account identifies several regions in the brain that may compose a ‘fatigue network,’ including the anterior insula, the anterior cingulate cortex and frontal/prefrontal areas ^12^. The exteroceptive and proprioceptive predictive coding account of fatigue predicts that in addition to the insula, networks comprised of sensorimotor, visual, and attentional hubs also contribute to fatigue ^15^. There is emerging evidence from resting-state functional magnetic resonance imaging studies that connectivity between these areas is associated with fatigue perception ^20–22^. Proprioception is impaired in individuals with perinatal stroke and is shown to be related to connectivity of sensory networks ^23,24^, supporting that the connectivity in these networks may contribute to fatigue. Following the recent suggestion that fatigue likely arises from *both* interoceptive and proprioceptive predictive processing ^17^, many of the hubs whose connectivity is associated with fatigue have also been suggested to be hubs within a global predictive processing network ^25–27^ At present, the relationship between fatigue in individuals with perinatal stroke and the functional connectivity of these networks has not been studied.

The aim of the present study was to examine the associations between fatigue and functional brain connectivity in individuals with perinatal stroke and compare this to the functional connectivity of typically developed youth. We have previously reported that corticospinal excitability was associated with fatigue severity in individuals with perinatal stroke ^6^. Thus, we hypothesized that the functional connectivity of the networks involved in proprioceptive and exteroceptive predictive processing would be associated with fatigue severity.

## Results

Seventy-five participants were identified through the Alberta Perinatal Stroke Project database. Of these, eleven were ineligible due to incomplete data, and four were removed due to excessive head motion in the scanner. The data for 60 stroke participants and 59 TDC were included in the analysis. Participant demographics and outcomes are summarized in Table 1. Lesion overlay maps for AIS participants are shown in Figure 1. The median age for the full sample was 11 years, range 7-19 years, and 35% were female. Forty-five percent (27/60) of the participants with perinatal stroke reported experiencing problems with fatigue (PEDSQL-CP fatigue score <u>≤</u>68.75) and were assigned to the “Fatigued” group ^6^ and the remaining 33 participants were assigned to the “Not Fatigued” group. There was no difference in age between the stroke groups. TDC were significantly older than the “Not Fatigued” group (see Table 1). There were no other differences in demographics between groups. (*p*>0.05).

**Table 1.** Demographics characteristics and fatigue scores

|  | Stroke<br>(N = 60) | Fatigued<br>(N = 27) | Not fatigued<br>(N = 33) | TDC<br>(N=59) |
| --- | --- | --- | --- | --- |
| Sex<br>(Female: Male) | 21:39 | 11:16 | 10:23 | 29:30 |
| Age<br>(years, mean±SD) | 11.5±2.8 | 11.5±2.5 | 11.5±3.1 | 14.2±3.0 |
| Stroke side<br>(Left: Right) | 37:23 | 19:13 | 18:10 | NA |
| Stroke type<br>(Arterial: Periventricular<br>Venous Infarction) | 29:31 | 15:12 | 17:16 | NA |
| Fatigue<br>(PEDSQL, median [IQR]) | 68.8 [32.8] | 50 [18.8] | 81.3 [25.0] | NA |

**Fig. 1.**
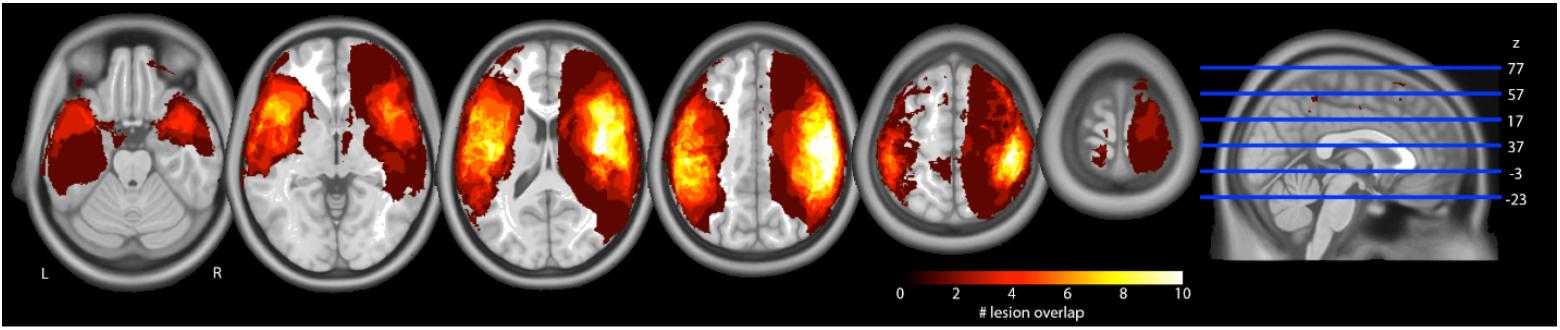
Lesion overlay maps for participants with arterial (AIS) strokes, shown are heat maps corresponding to the number of patients that have a lesion in that area overlaid on axial slices from a standard template in MNI space (MNI152). White shading indicates most lesion overlap and dark red least overlap. Images are presented in neurological convention (i.e., right hemisphere is on the right side).

Fisher-transformed correlation coefficients for ROI-to-ROI connections, age, PSOM score and stroke type (AIS or PVI) were entered into the model. The model selected three FC values: FC between the non-lesioned supramarginal gyrus and middle frontal gyrus, the non-lesioned insula and lesioned inferior temporal gyrus, and the lesioned paracingulate cortex and non-lesioned intracalcarine cortex. These connections are shown in Figure 2.

**Figure 2.**
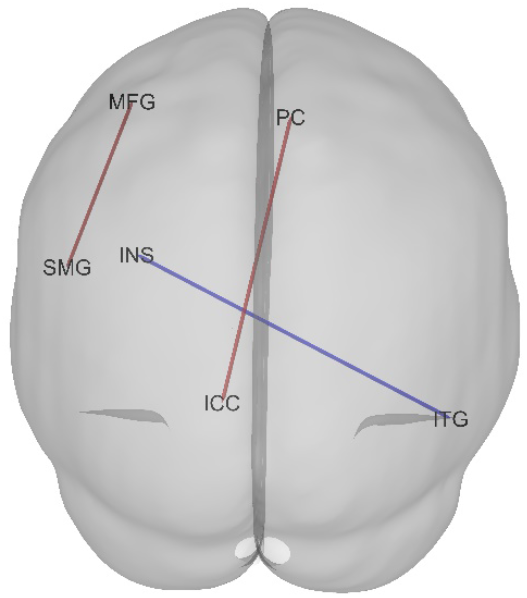
FC identified by the 10-fold cross-validated elastic net regression. Red lines illustrate a positive correlation, where a lower PEDSQL fatigue score (indicating more severe fatigue) was associated with weaker FC. The lesioned hemisphere is displayed on the left of this figure. MFG; middle frontal gyrus, SMG; supramarginal gyrus, ICC; intracalcarine cortex, PC; paracingulate gyrus, INS; insula cortex, ITG; Inferior temporal gyrus.

### Associations between Fatigue and FC

While associations between fatigue and FC were strong (as identified previously), the directionality of such associations differed by ROI-ROI pair. In participants with perinatal stroke, fatigue (PEDSQL score) was positively associated with FC between the non-lesioned middle frontal gyrus and supramarginal gyrus (β=19.6, SE=6.4, p=0.002, Figure 3A) and between the lesioned paracingulate gyrus and the non-lesioned intracalcarine cortex (β=31.6, SE=12.6, p=0.015, Figure 3C). In contrast, fatigue was negatively associated with FC between the non-lesioned insula and the lesioned inferior temporal gyrus (β=−42.3, SE=7.7, p<0.001, Figure 3B). Age, stroke type or sensorimotor function (PSOM) were not identified as predictors.

**Figure 3.**
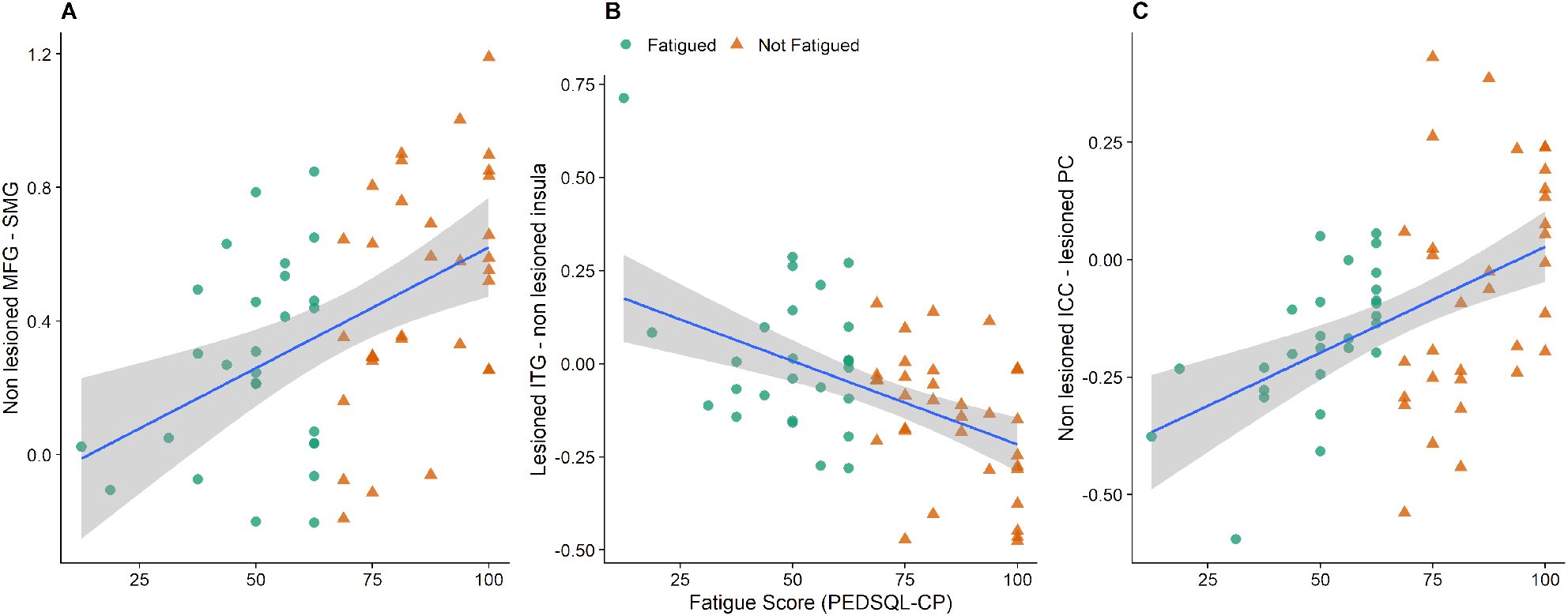
Associations between FC identified by the elastic net regression and fatigue (PEDSQL fatigue score). A lower fatigue score represents more severe fatigue. MFG; middle frontal gyrus, SMG; supramarginal gyrus, ICC; intracalcarine cortex, PC; paracingulate gyrus, ITG; Inferior temporal gyrus.

### Differences in functional connectivity between groups

Results from the Welch’s ANOVA for the Fisher-transformed correlations selected by the elastic net regression for fatigued, not fatigued and TDC participants are shown in Table 2 and Figure 4. Welch’s ANOVA revealed a difference between groups for all three FC ROI pairs. FC between the non-lesioned middle frontal gyrus and supramarginal gyrus was stronger in not fatigued participants compared to fatigued participants (*p*_*(fdr)*_=0.015, *d*=0.67), and TDC (*p*_*(fdr)*_*=0*.*002, d*=1.88), whilst it was stronger in fatigued participants than in TDC (*p*_*(fdr)*_=0.002, *d*=1.14). The negative FC between the non-lesioned insula and lesioned temporal gyrus was stronger in not fatigued participants compared to fatigued participants (*p*_*(fdr)*_=0.002, *d*=0.88) and TDC (*p*_*(fdr)*_=0.002, *d*=1.12) but not different between fatigued participants and TDC (*p*_*(fdr)*_=0.273, *d*=0.35). The negative FC between the non-lesioned intracalcarine cortex and the lesioned paracingulate cortex was stronger in fatigued participants compared to not fatigued (*p*_*(fdr)*_=0.045, *d*=0.56), and TDC participants (*p*_*(fdr)*_=0.002, *d*=0.94), and not different between not fatigued and TDC participants (*p*_*(fdr)*_=0.149, *d*=0.30).

**Table 2.** Means + SD for Fisher-transformed correlations for the ROI-to-ROI analysis

|  | Fatigued<br>(N = 27) | Not Fatigued<br>(N = 33) | TDC<br>(N = 59) | Group differences<br>(Welch's ANOVA) |
| --- | --- | --- | --- | --- |
| Non-lesioned (left) middle frontal gyrus –<br>supramarginal gyrus (posterior) | 0.27±0.30 | 0.50±0.34 | -0.03±0.23 | $F_{(2,51)}=32.6, p<0.001$ |
| Non-lesioned (left) insula – lesioned (right) inferior<br>temporal gyrus | 0.02±0.21 | -0.15±0.18 | 0.10±0.25 | $F_{(2,65)}=16.2, p<0.001$ |
| Non-lesioned intracalcarine cortex – lesioned<br>paracingulate cortex | -0.17±0.15 | -0.06±0.24 | 0.02±0.21 | $F_{(2,65)}=10.8, p<0.001$ |

In TDC; lesioned = right, non=lesioned = left, TDC; typically developed control participants

**Figure 4.**
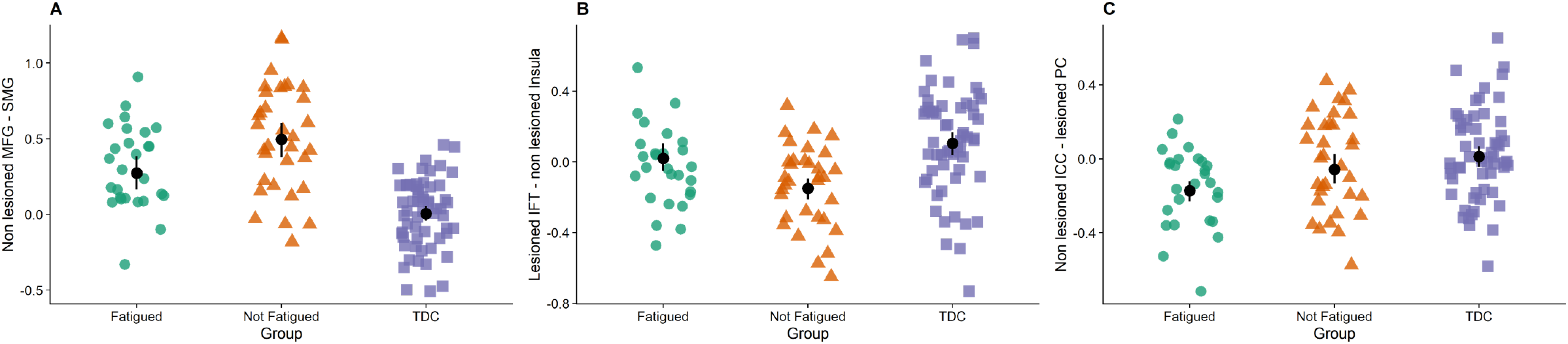
Fisher-transformed correlation coefficients for the connectivity between the A) non-lesioned primary middle frontal gyrus (MFG) and supramarginal gyrus (SMG), B) lesioned inferior temporal gyrus (ITG) and non-lesioned insula and C) between the non-lesioned intracalcarine cortex (ICC) and lesioned paracingulate cortex (PC). The black circle and error bars represent the mean±95% confidence interval. TDC; Typically developed controls.* represents a different (p<0.05) FC compared to the not fatigued group.

## Discussion

The aim of the present study was to examine potential associations between fatigue and resting-state functional connectivity in the brains of individuals with perinatal stroke. Compared to participants who were not fatigued, those who reported problems with fatigue had weaker connectivity between the non-lesioned middle frontal gyrus and posterior supramarginal gyrus, and between the non-lesioned intracalcarine cortex and lesioned paracingulate gyrus. In contrast, functional connectivity between the non-lesioned anterior insula cortex and the lesioned inferior temporal gyrus was stronger in individuals experiencing fatigue. This study is the first to explicitly test the relationship between functional brain connectivity and perceived fatigue in young people with perinatal stroke. These results suggest that fatigue is associated with the functional connectivity of the interoceptive and salience prediction networks in individuals with perinatal stroke.

Here, we provide evidence showing that fatigue in perinatal stroke is associated with altered connectivity between hubs in previously identified global predictive coding networks ^25–27^. These include hubs frequently linked to proprioceptive and interoceptive processing and allostatic control ^28–30^. Within predictive coding frameworks, fatigue is suggested to arise from an ongoing and unresolvable discrepancy between the brain’s prediction of the body-state and the afferent sensory information signalling the actual-body state, the prediction errors ^11,12,14^. Unresolvable prediction errors signal a failure for the brain to predict the body state, i.e. an inability to maintain allostatic control, consciously perceived as the feeling of fatigue ^12^. We found that individuals experiencing more fatigue had weaker FC between the non-lesioned supramarginal gyrus and middle frontal gyrus compared to those who did not experience fatigue. Recent evidence suggests that these areas are structurally connected via the superior longitudinal fasciculus ^31^. Perhaps surprisingly, the weaker connectivity seen in fatigued participants reflected the pattern in typically developed participants, where there was virtually no functional connectivity between these areas. In adults and children with stroke, the supramarginal gyrus and middle frontal gyrus have been related to successful proprioceptive processing ^32–34^, while individuals with perinatal stroke may have impaired proprioceptive function^23,24^. Within the predictive processing framework, both areas are associated with both prediction formation and error ^26,27^. When comparing participants who did not experience fatigue with TDC, we speculate that stronger functional connectivity between these areas seen in not fatigued participants represents possible compensatory neural plastic alterations related to impaired proprioceptive prediction and prediction error processing. Kuppuswamy ^11^ suggests that fatigue arises from dysfunction in the systems involved in processing proprioceptive prediction errors, suggesting that the weaker functional connectivity, or an inefficient compensation mechanism, between these areas may be related to the increased perceptions of fatigue.

A similar pattern can be observed in the functional connectivity between lesioned inferior temporal gyrus and non-lesioned insula, wherein not fatigued participants had stronger (negative) functional connectivity than fatigued participants, who more closely resembled typically developed participants. The insula, cingulate and inferior temporal gyrus are suggested to be hubs within a neural network that supports interoceptive processing and allostasis ^30^. An alternative predictive processing account of fatigue suggests that fatigue arises from continued and unresolvable interoceptive, not proprioceptive, prediction errors which signal failures in allostatic control ^12^. Again, when interpreted through the predictive processing theoretical framework, the stronger connectivity seen in not fatigued participants suggests adaptive plasticity supporting interoceptive predictive processing. The present data indicate the involvement of both proprioceptive and interoceptive predictive processing in the development of fatigue in individuals with perinatal stroke. Although the proprioceptive and interoceptive predictive processing accounts of fatigue are often contrasted ^14,15^, it is more likely that fatigue can arise from ongoing and unreconcilable proprioceptive and interoceptive prediction errors, which both signal challenges to allostatic control ^17^. Since fatigue is believed to be the affective response signalling ongoing unresolvable prediction errors and thus failure to exert allostatic control, these results support the predictive processing accounts of fatigue ^14,15^. At present, there is considerable uncertainty about how predictive processing networks function and much more evidence of the strength and direction of relationships is required before this suggestion can be genuinely considered.

Somewhat surprisingly, and in contrast to our hypothesis, we did not find any associations between fatigue and the connectivity between primary or secondary sensorimotor areas. This is in contrast to an earlier analysis of this data, where we used a theory-driven approach and placed seeds within these areas ^35^, as well as our previous study where we used transcranial magnetic stimulation to probe the association between fatigue and corticomotor excitability ^6^ and previous evidence of altered FC within the sensorimotor network associated with motor function ^36–38^. However, this is not to say that functional connectivity within sensorimotor areas is not associated with fatigue, indeed we have shown that it is ^29^. Instead, what the current data show is that of more relevance to the perception of fatigue in individuals with perinatal stroke is the connectivity of hubs within predictive processing networks.

This study has some limitations which should be acknowledged. We have interpreted our results using the predictive coding theories of the brain and affect. While our data support these models, we did not measure the activity of this network during an action ^39^, nor did we have a measure of predictive coding during performance ^40^. These are the necessary next steps in examining fatigue in youth with perinatal stroke. Although we applied post-hoc corrections for head movement, fMRI remains a challenging technique in a pediatric population, and we were required to remove several participants from our sample. The replication of these effects in adults with perinatal stroke may overcome this limitation and provide a method to replicate and extend these results. Our sample was composed of participants capable of completing an MRI, which is necessarily the less disabled and higher functioning portion of the perinatal stroke population.

## Conclusion

We found that fatigue in individuals with perinatal stroke was associated with altered functional connectivity of a global predictive processing network. These findings provide interesting avenues for both future research and therapeutic interventions. The effects of emerging therapies, such as novel training paradigms and non-invasive brain stimulation, on both neural functional connectivity and fatigue are warranted.

## Method and Materials

### Participants

Participants were recruited via the Alberta Perinatal Stroke Project ^41^ and the Healthy Infants and Children Clinical Research Program (www.hiccupkids.ca) research cohorts at the Alberta Children’s Hospital. Inclusion criteria for cases were age 6-19 years; term birth; magnetic resonance imaging (MRI)-confirmed unilateral perinatal arterial ischemic stroke (AIS) or periventricular venous infarction (PVI), symptomatic hemiparetic cerebral palsy including Pediatric Stroke Outcome Measure >0.5; Manual Ability Classification Scale levels I-IV. Inclusion criteria for typically developed controls (TDC) were right-handed, ages 6-19 years, and no neurological conditions, medications, or MRI contraindications. Participants and parents provided written informed assent/consent. Experiments were carried out with the approval of the Research Ethics Board at the University of Calgary and in accordance with the declaration of Helsinki ^42^.

### Fatigue

Self-reported fatigue was assessed in participants with perinatal stroke using the fatigue subscale of the Pediatric Quality of Life Inventory 3.0 cerebral palsy module (PEDSQL-CP), a validated measure of quality of life in children with CP ^43^. The fatigue subscale consists of four statements: “I feel tired,” “I feel physically weak (not strong),” “I rest a lot,” and “I don’t have enough energy to do things that I like to do.” Individuals were asked to give a rating of 0-4 for each statement if they never (0), almost never (1), sometimes (2), often (3), or almost always (4) experienced a problem with the four areas of fatigue highlighted in the statements. Parents assisted children under eight years with the rating using the PEDSQL 3.0 CP module parent report for young children. The PEDSQL scores for fatigue were reverse scored and linearly transformed to a 0 to 100 scale ^43^. The average score across all categories in each subscale was calculated. Lower scores in each subscale represent higher levels of fatigue. Participants were classified as experiencing fatigue if they scored ≤68.75, indicating a problem with at least two or more of the categories of fatigue, or if they scored 4 (“almost always” a problem) for one of the categories ^5^.

### MRI collection

MRI sequences were acquired according to a standardized perinatal stroke neuroplasticity protocol at the Alberta Children’s Hospital Diagnostic Imaging Suite using a 3.0 Tesla GE MR750w MRI scanner (GE Healthcare, Waukesha, WI) with an MR Instruments 32-channel head coil. High-resolution T1-weighted anatomical images were acquired in the axial plane [166 contiguous slices; voxel size = 1.0 mm isotropic; repetition time (TR) = 8.5 ms; echo time (TE) = 3.2 ms]. Resting-state fMRI acquisition used 150 T2*-weighted whole-brain echo-planar volumes (EPI; 36 interleaved contiguous slices; voxel size = 3.6 mm isotropic; TR/TE = 2000/30 ms). Participants were told to fixate on a centrally presented cross. Images were reoriented such that the stroke was located in the right hemisphere for all patients so that lesioned hemispheres of cases corresponded to non-dominant hemispheres in controls.

### Functional connectivity analyses

Resting-state functional connectivity analyses were performed using the SPM12 (Statistical Parametric Mapping, Wellcome Trust, UCL, UK) Functional Connectivity Toolbox (CONN) ^44^ in Matlab version r2018a (Mathworks, Natick, MA, USA). Slice timing correction, realignment, and co-registration were performed, and head motion parameters were estimated. Co-registered images were segmented using subject-specific tissue probability maps. Images were normalized into Montreal Neurological Institute (MNI) space via direct non-linear transformations using the standard 152-average template. Direct normalization results in structural and functional volumes being separately normalized to MNI space. Images were then smoothed with a 6 mm^3^ full-width at half-maximum Gaussian kernel. Head motion exceeding 0.9 mm of translational head movement and global intensity outliers (z-score>5) were identified using the Artifact Repair Toolbox ^45^. Time courses of blood oxygenation level-dependent (BOLD) responses were extracted for the GM, CSF and WM. The CSF, WM and lesion time courses were regressed from the general linear regression model (GLM). Outlier volumes and head motion were additionally de-weighted in the GLM.

In a preliminary analysis of this data ^35^, we performed both ROI-to-ROI and seed to voxel analysis in eight predetermined ROI’s within the sensorimotor network. However, based on feedback to this preprinted analysis, and subsequent experimental and theoretical advances in the study of fatigue ^15,21,22^, we decided to adopt a data-driven analysis plan to reduce possible bias in seed selection.

Individuals with perinatal stroke often have widespread morphological and functional differences in brain structure and development (^46^ for a review). Because of these differences, we could not perform component identification whole-brain connectivity analyses (e.g., independent component analysis, principal component analysis). Instead, we used a machine learning ROI-to-ROI approach. All ROIs defined using the default Harvard-Oxford Atlas within CONN were included in the analysis, except those in the cerebellum and vermis. To reduce the number of FC, the two ROIs for the cerebellum were selected from the CONN ICA analysis of the Human Connectome Project dataset ^44^, resulting in 108 ROIs entered into the analysis. BOLD signal time series were calculated using the average of all voxels within the ROI. Fisher-transformed correlation coefficients were interpreted as quantifications of functional connectivity (FC) strength between areas. FC between each of the ROIs were identified in CONN and extracted for data analysis using elastic net regression. FC values between ROIs identified as associated with fatigue in children with perinatal stroke using the elastic-net regression model (described below) were subsequently quantified in TDC to compare functional connectivity between participant groups.

### Sensorimotor function

We included a measure of sensorimotor function to quantify whether function mediated the relationship between fatigue and the connectivity of the sensorimotor prediction network. Sensorimotor function was assessed via subscales of the Pediatric Stroke Outcome Measure (PSOM). The score for the more affected hand was used. The PSOM is a reliable and valid measure of neurological function following stroke in children ^47^ and the sensorimotor subscale evaluations include motor, visual, hearing, and somatosensory function ^48^.

### Data and statistical analysis

Statistical analyses were performed with the R statistical software package ^49^. Analysis of variance (ANOVA), chi-square and Mann-Whitney U tests were used to compare the demographic data. To identify FC between ROIs that was associated with fatigue, stroke participants’ Fisher-transformed coefficients (N = 5778 ROI-to-ROI connections), demeaned age, PSOM score and stroke type (AIS or PVI infarction) were entered into an elastic net regularized regression model, with fatigue score as the outcome variable. Ten-fold cross-validated elastic net regression was performed using the ‘glmnet’ package ^50^. Elastic net regression is a regularized regression technique, used for variable selection in high dimension low N studies, which uses a penalty term to shrink regression coefficients of multiple predictors, setting some to 0 and leaving only the most relevant predictors in the model. A subsequent robust linear regression model containing predictors identified in the model was used to determine model goodness-of-fit and estimate coefficients. Connectivity identified in the elastic net model was compared between groups (Fatigued, Not fatigued, TDC) using Welch’s analysis of variance (ANOVA). Significant main effects were followed up using *post hoc* Welch’s t-tests. Shapiro-Wilk’s test of normality was used to assess the distribution of the residuals. Cohen’s *d* and its associated 95% confidence were used as an estimate of the effect sizes for differences in connectivity between groups. The threshold to reject the null hypothesis was set at *p*<0.05. Where necessary, the false-discovery rate was adjusted to control for multiple comparisons ^51^.

## Data Availability

Data is available upon reasonable request from the corresponding author

## Data availability statement

Raw data is available upon reasonable request from the corresponding author. Code used for analysis is availble from the Open Science Framework (https://osf.io/u4jvh).

## Author Contributions

Author contributions included conception and study design (JGW, HC, EGC, AK) data collection and acquisition (HC, AK) statistical analysis (JGW), interpretation of results (JGW, HC, EGC, AK), drafting the manuscript work or revising it critically for important intellectual content (JGW, HC, EGC, AK)) and approval of final version to be published and agreement to be accountable for the integrity and accuracy of all aspects of the work (All authors).

## Compliance with Ethical Standards

Experiments were carried out with the approval of the Research Ethics Board at the University of Calgary and in accordance with the declaration of Helsinki ^42^.

## Supplementary material

The following is a description of the results from the first iteration of this study, where the analysis of the association between fatigue core and ROI-ROI FC was constrained to the FC between sensorimotor areas “The model identified two ROI-to-ROI connections as significant predictors of fatigue status. FC between the non-lesioned S1 and lesioned SMA (β= −21.7, SE = 9.9, p = 0.032) and the lesioned and non-lesioned thalamus (β= 15.5, SE = 7.5, p = 0.032) were significant predictors of fatigue score (R2 = 0.15, F(2,57) = 4.9, p = 0.017, Figure 2).

